# New insights into infant strongyloidiasis in Papua New Guinea

**DOI:** 10.1101/2025.04.17.25325968

**Authors:** Huan Zhao, Juciliane Haidamak, Eva Noskova, Vladislav Ilik, Barbora Pafčo, Rebecca Ford, Geraldine Masiria, Tobias Maure, Nichola Kotale, William Pomat, Catherine Gordon, Severine Navarro, Paul F. Horwood, Constantin Constantinoiu, Andrew R. Greenhill, Richard S. Bradbury

## Abstract

The island of New Guinea is known to harbour a unique human infecting species of *Strongyloides*: *Strongyloides fuelleborni* subsp. *kellyi*. Stool DNA extracts (n=164) from 19 PNG infants, collected over 13 months, were analysed using *Strongyloides* real-time PCRs and underwent metabarcoding of *cox1*, *18S rRNA* HVR-I and HVR-IV loci. Eight infants were infected with *Strongyloides*; seven identified as *Strongyloides fuelleborni fuelleborni* and one as *S. f. kellyi*. Phylogenetic and haplotyping analyses indicated that *S. f. fuelleborni* in PNG belongs to the Indochina sub-clade of *S. f. fuelleborni,* and *S. f. kellyi* does not represent a subspecies of *S. fuelleborni*.

We provide the first molecular evidence of *S. f. fuelleborni* infection in humans in the Pacific. We also support that *S. f. kellyi* is a separate and distinct species of human infecting helminth. Renewed clinical and epidemiological investigations into infant strongyloidiasis in New Guinea using species-discriminatory molecular tools are indicated.

## Introduction

New Guinea, consisting of Indonesian New Guinea in the west and Papua New Guinea (PNG) on the east, is the world’s largest tropical island and a biodiversity hotspot. The island is known to harbour a unique and enigmatic agent of human strongyloidiasis, *Strongyloides fuelleborni* subsp. *kellyi*, along with the more widespread and better understood human pathogen, *Strongyloides stercoralis*(1–3). This *Strongyloides fuelleborni*-like intestinal nematode of humans was first reported by Kelly and Vogue(4) in western PNG in 1971 and later found by Muller et al.(5) in Indonesian New Guinea.

Taxonomic confusion over the identity of this New Guinea *Strongyloides* and its relationship to African *S. fuelleborni* subsp. *fuelleborni* soon arose. Unlike *S. stercoralis*, which is passed as rhabditiform larvae, *S. f. kellyi* is passed as eggs, resembling those of larvated hookworms, in faeces(1, 6). Viney et al.(3) observed that the adult nematodes were morphologically distinguishable by subtle differences in the peri-vulval cuticle of parasitic females and the phasmidial pore position of free-living males(3). A separate isoenzyme electrophoresis analysis indicated that both worms clustered closely, separate from other mammal *Strongyloides* spp.(7). Based on these findings, Viney et al.(3) proposed subspeciation of *S. fuelleborni* von Linstow, 1905 into *S. f. fuelleborni* and *S. f. kellyi*, the latter designating the New Guinea nematode. Notably, Asian strains of *S. fuelleborni* were not included in the analyses by Viney et al.(3).

The epidemiology of *S. f. kellyi* is similarly enigmatic. Patent infection in children has been observed within 3 weeks after birth(6). An intervention study found that the incidence of infection rose rapidly in the first 12 months of life, stabilising thereafter until 5 years, at which age incidence began to drop(8). How infection occurs in children so young is unclear. Transmammary transmission has been suggested(6). Screening of breastmilk from 40 mothers in an endemic village during the 1970s revealed no larvae(9). Non-human primates (NHPs), the animal reservoir of *S. f. fuelleborni*, do not occur in New Guinea and attempts to find an animal reservoir for *S. f. kellyi* by screening domestic animals, including pigs, chickens and dogs from villages where infections occurred at high prevalence in humans were unsuccessful(4, 7).

Heavy infection with *S. f. kellyi* has been implicated as the cause of a rapidly fatal disease in infants around 2 months of age, known as “swollen belly syndrome” (SBS)(6). SBS is a protein-losing enteropathy characterised by eosinophilia, distended abdomen, diarrhoea, and respiratory distress(10). The aetiology of SBS remains poorly understood. This disease was observed between 1974 and 1983, predominantly in two remote villages in mountainous regions of PNG, Kanabea (Gulf Province, ∼1270 metres above sea-level) and Wanuma (Madang Province, ∼750 metres above sea-level). Only occasional, sporadic, cases were reported elsewhere(6). In Kanabea, patent infection with an egg producing *Strongyloides* sp. was found in 100% of infants 3-8 weeks after birth, with a mean average egg burden of 94,000 eggs/mL(9). SBS caused 8% of infant deaths in this village until specific anthelmintic therapy was introduced(6). Since the early 1980s, we are only aware of one identified case of SBS in PNG, occurring circa 2010 (Andrew Greenhill, pers. comm. 2024). This is despite the very high burdens and prevalence of *Strongyloides* eggs, assumed to represent *S. f. kellyi*, demonstrated in infants in several other parts of PNG, where it is associated with nutritional deficiencies, but not SBS(11, 12). Clearly another unknown factor was involved in the development of SBS(10).

Dorris et al.(13) provided the only *S. f. kellyi* DNA sequence to date, a 330bp segment of *18S* (syn. SSU) *rRNA*, which covers the hypervariable region (HVR)-I(14). Phylogenetic analysis based on this region placed *S. f. kellyi* in a clade with *Strongyloides cebus*, *Strongyloides papillosus*, and *Strongyloides venezuelensis*, but distant from *S. f. fuelleborni*. This work prompted calls to elevate *S. f. kellyi* to the species rank(2). However, a criticism of this finding is that *18S* HVR-I has been indicated as a poor marker for inferring taxonomic positions of *Strongyloides* spp. (15, 16). This gene region did not allow the separation of *Strongyloides* spp. with spiralled and straight ovary morphotypes, whereas nearly full-length *18S rRNA* and *28S rRNA* sequences did(13, 17). Furthermore, Dorris et al.(13) found *S. f. fuelleborni* to be phylogenetically closer to *S. stercoralis* than to *S. papillosus,* contradicting evidence suggested by mitochondrial genome data(18). Molecular taxonomy of *Strongyloides* spp. determined using markers such as mitochondrial cytochrome oxidase subunit 1 (*cox*1) and *18S* HVR-IV showed considerable consistency with whole-genome and mitochondrial genome analyses(18, 19). The true phylogeny and taxonomic identity of *S. f. kellyi* needs to be elucidated using these informative markers.

Despite its public health significance, *S. f. kellyi* remains a neglected and underexplored human helminth. This enigmatic causative agent of strongyloidiasis in New Guinea warrants focused research to clarify its true identity and epidemiology.

## Methods

### Sampling

Samples were obtained as part of a 1-year longitudinal study investigating gut health in infants in Eastern Highlands Province, PNG, during 2018-2019 (Figure 1). Ethics approval was obtained from the Papua New Guinea Institute of Medical Research Institutional Review Board (IRB# 1614) and the Papua New Guinea Medical Research Advisory Committee (approval no. 17.17). Infants were enrolled into the study within the first 5 weeks of life. Initially parents assented to participate into the study during antenatal clinic visits. After giving birth, informed consent was obtained from at least one parent or legal guardian. Stools samples were sought from participants monthly for 12 months. As part of this study, we investigated the presence of *Strongyloides* spp. in infant stool samples in 19 infants, 12 males and 7 females. For samples used in the *Strongyloides* spp. study, infants were aged between 0 and 101 days (average: 30 days) when they provided their first stool sample. Clinical data on the participants was limited, but no marked gastrointestinal pathology was noted during the course of the study.

**Figure 1.**
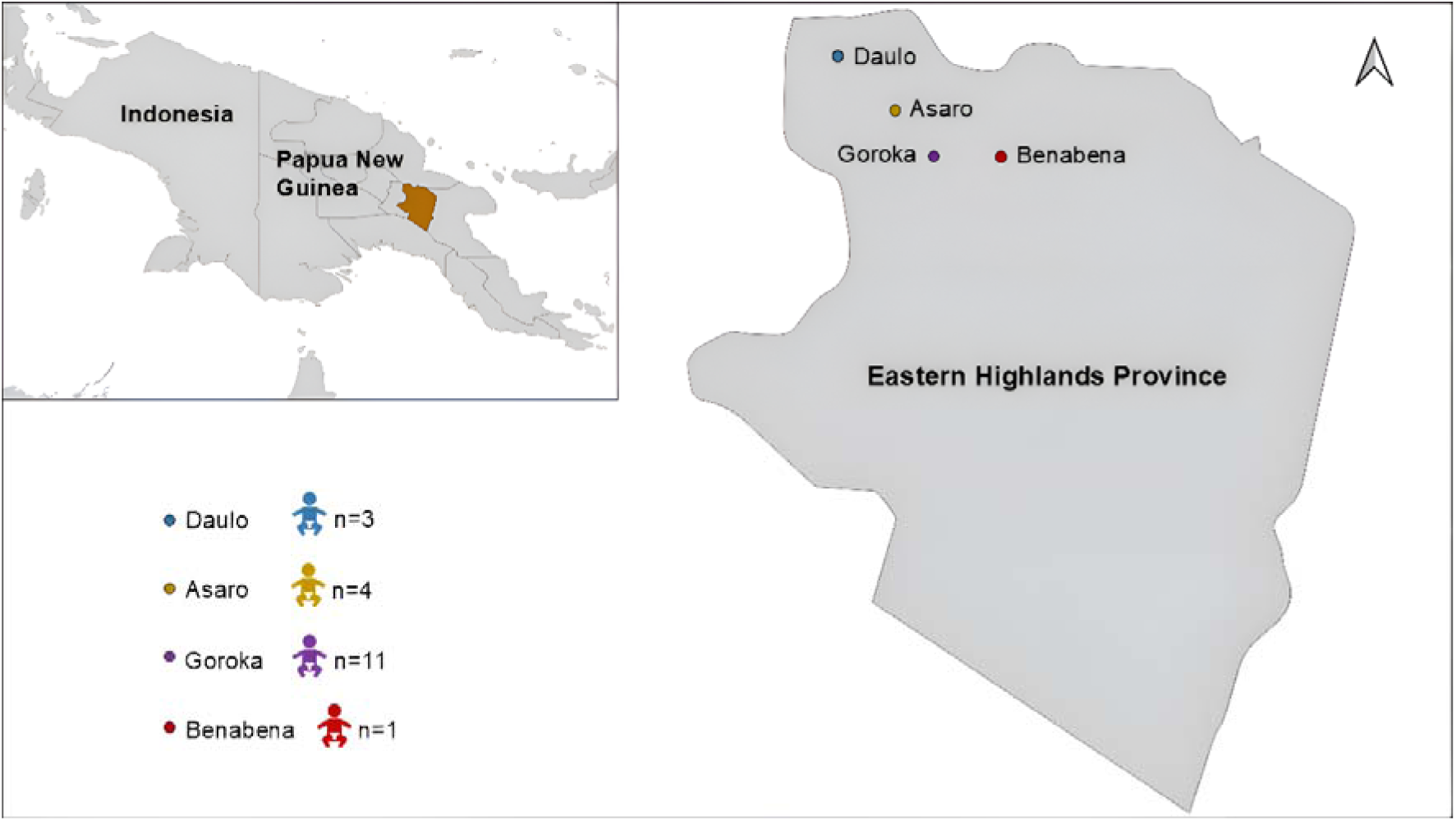
Sampling locations, with participant numbers from each location, from Eastern Highlands Province, Papua New Guinea.

Fresh stool samples were transported to the PNG Institute of Medical Research (Goroka, PNG), where they were then stored at -70 °C. Samples were subsequently sent to QIMR Berghofer Medical Research Institute (Queensland, Australia) and James Cook University (Queensland, Australia) for laboratory analysis. We extracted DNA from each sample using QIAamp DNA mini kit (Qiagen, Hilden, Germany), following the manufacturer’s protocol with the following pre-extraction steps: Stool samples (∼200mg) were mixed with 500µL of ROSE buffer, 15µL of proteinase K, and 500µL of 0.5mm silica/zirconia beads (Daintree Scientific, St Helens, Australia) in 2mL screw-cap tubes (Starstedt). The tubes were homogenised at 6500 rpm for 40 seconds using a Precellys homogeniser. Following homogenisation, the samples were incubated at 95°C for 5 minutes and then centrifuged at 4000g for 3 minutes. The supernatant was transferred to a new 1.5 mL tube, and the manufacturer’s protocol was conducted from this point.

Extracted faecal DNA was assessed for quality and quantity using a NanoDrop 2000. DNA samples with a concentration greater than 10 ng/µl, and a 260/280 ratio between 1.6 and 2.10 were used as qPCR templates. Samples that did not meet those requirements were re-extracted. A total of five samples were re-extracted, and all subsequently reached the required DNA concentration. DNA was then diluted 1:5 with MilliQ water and subjected to two published *Strongyloides* real-time PCR (qPCR) assays(20, 21). Positive qPCR results were confirmed by triplicate testing and were considered positive when the cycle threshold (Ct) was below 35. To tentatively infer the identity of the qPCR product, Sanger sequencing of positive amplicons by the two qPCRs was performed, targeting a 471 bp region of *18S rRNA*(20) and a 138 bp repeat sequence(21) respectively, using the BigDye Direct Cycle Sequencing Kit (Thermo Fisher Scientific, Waltham, USA).

We also performed a metabarcoding assay targeting *18S rRNA* HVR-IV (∼252bp) using the Illumina MiSeq Reagent Nano Kit v2 (Illumina, San Diego, CA)(16). Positive samples were subsequently subjected to additional metabarcoding of the *18S rRNA* HVR-I (∼432bp) and partial *cox*1 (217bp) genotyping targets(16) using the same Illumina MiSeq platform. Sequencing was conducted with 500 cycles (PE250bp) for *18S rRNA* products and 300 cycles (PE150bp) for the *cox*1 products to ensure adequate sequencing coverage.

### Statistical and bioinformatic analysis

Demographic and qPCR data were imported into Microsoft Excel for statistical analysis. Raw Illumina sequencing data were independently analysed by three blinded researchers at James Cook University (JCU) and the Institute of Vertebrate Biology (IVB) using two different computational pipelines: a custom Geneious Prime (https://www.geneious.com/) workflow following Barratt et al.(16) (JCU and IVB) and a combination of Skewer(22) and DADA2(23) packages implemented in R software (v4.2.2) (https://www.r-project.org/) (IVB). Both pipelines incorporated read quality control, contig assembly, and haplotype assignment. Maximum likelihood and Bayesian inference phylogenetic analyses were performed on MUSCLE-aligned *cox1* sequences, using the GTR model for nucleotide substitutions, in MEGA 11 and MrBayes respectively. Cohen’s kappa was then used to assess the diagnostic agreement between different molecular methods for the detection of *Strongyloides* spp.

## Results

A total of 164 stool samples were collected from 19 infants across five to 13 occasions over the study period (Figure 1 & 2). Eight of 19 infants (42%), comprising six males and two females, were found to be infected with a *Strongyloides* sp. infections were detected at an average age of 212 days (range: 94-334 days) (Figure 2). Of the eight positive cases, faecal metabarcoding at *cox*1, *18S rRNA* HVR-I and HVR-IV loci identified six infections as *S. f. fuelleborni* and one as *S. f. kellyi* (Table 1). The remaining one positive sample could only be amplified at the qPCR *18S rRNA* target. This 471bp sequence was 99.2% identical to a published *S. f. fuelleborni* sequence by BLAST analysis (GenBank accession: AB272235).

**Figure 2.**
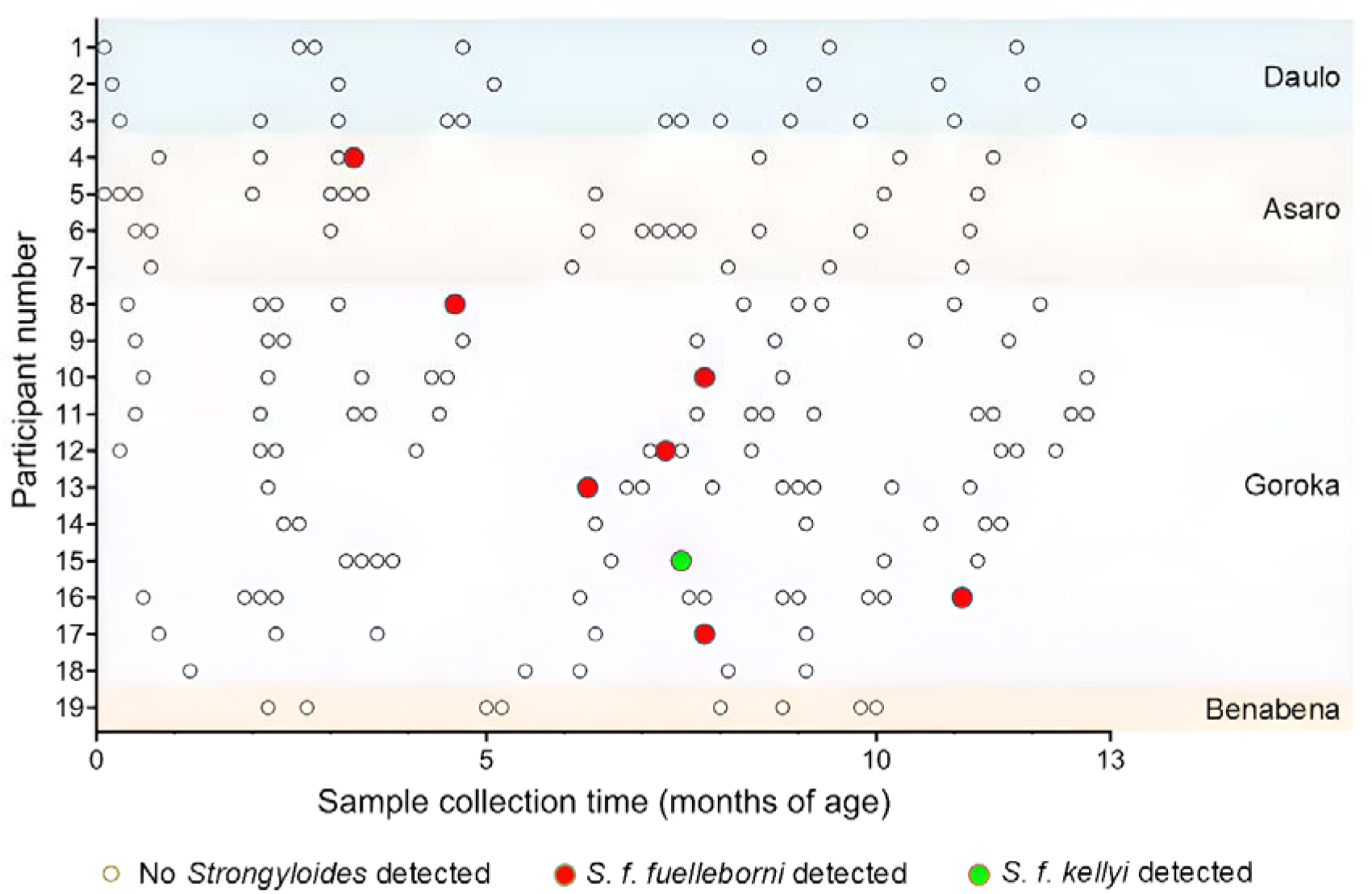
Sampling frequency and *Strongyloides* infection in infants from Eastern Highlands Province, PNG. Multiple samples (at least five) were collected from each infant enrolled over the first 13 months of life. Samples which demonstrated molecular evidence of *Strongyloides* spp. infection are shown.

**Table 1.**
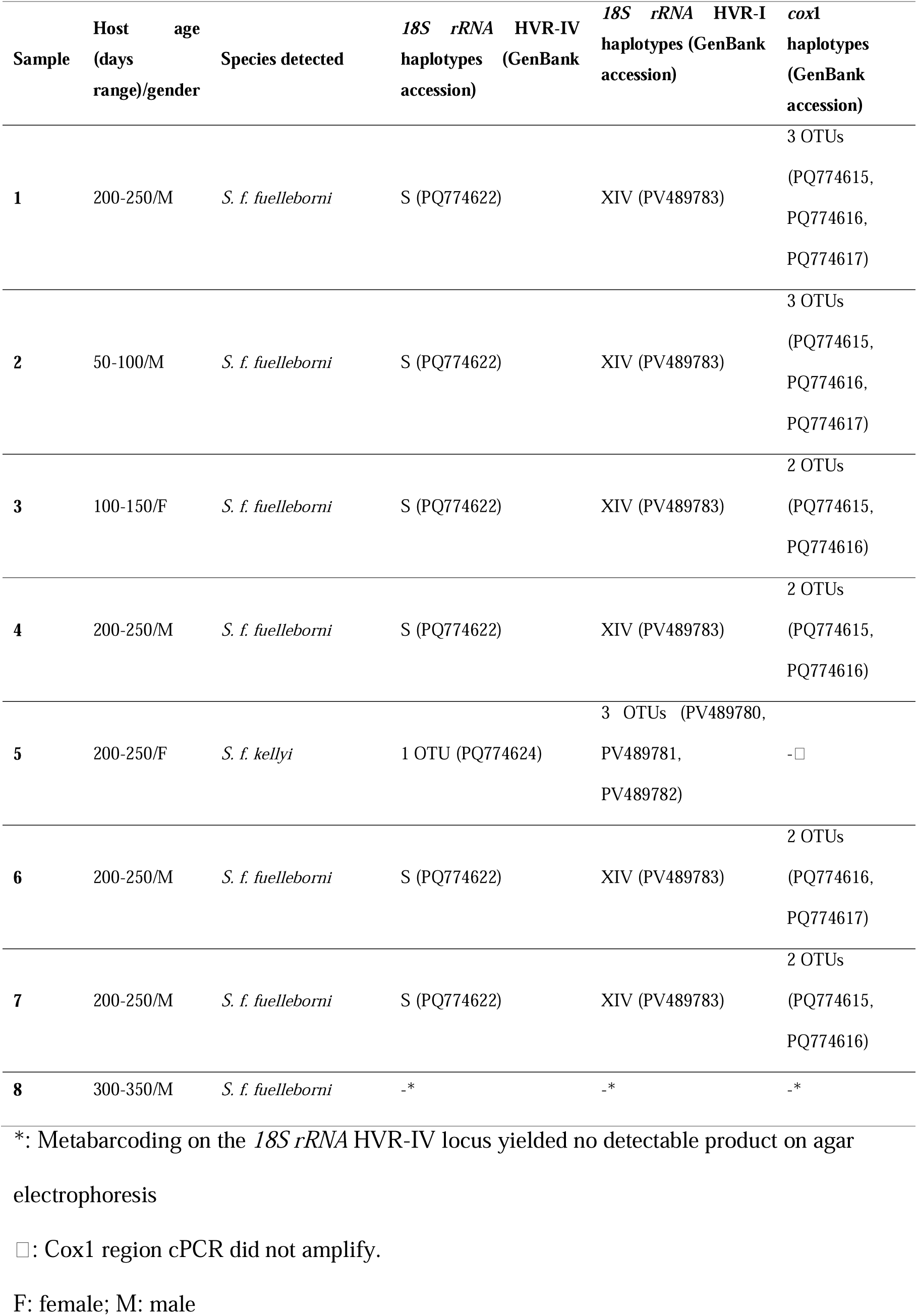
*Strongyloides* samples analysed in this study and their genotypes.

The genus-specific qPCR of Llewellyn et al. (20) performed comparably with the Barratt et al. (16) *18S rRNA* HVR-IV metabarcoding in detecting *Strongyloides* infections, with a Cohen’s kappa coefficient of 0.85. The Pilotte et al. (21) qPCR only detected *S. f. kellyi* (Figure 3).

**Figure 3.**
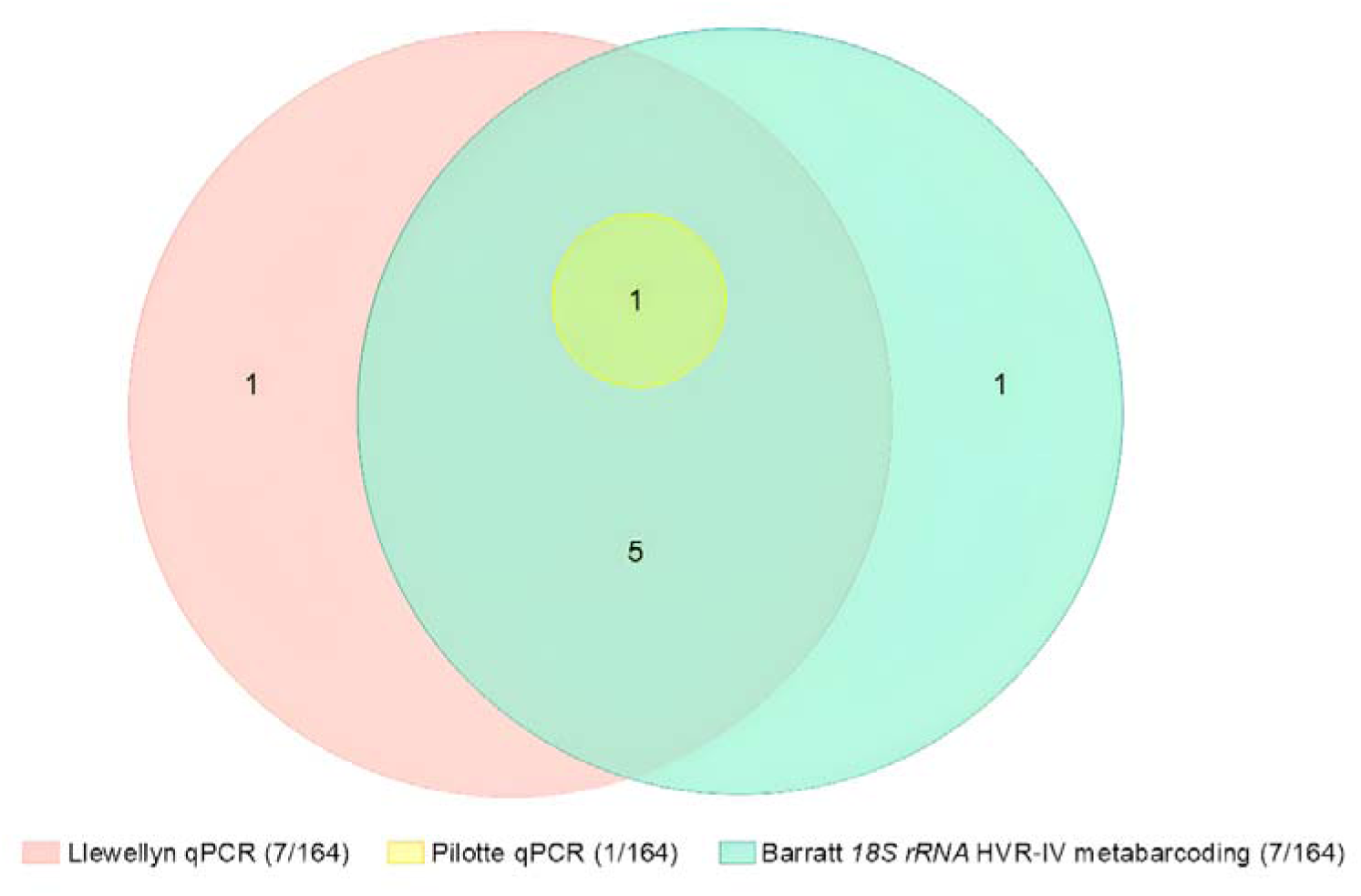
Euler diagram showing the performance of two real-time PCRs (qPCR)(20, 21) and *18S rRNA* HVR-IV metabarcoding(16) for the detection of *Strongyloides* spp. in 164 infant stool samples from Papua New Guinea.

Sequences of *18S rRNA* HVR-I (432bp) and HVR-IV (∼252bp) were obtained from seven samples (**Table 1**). Six positive samples harboured HVR-IV haplotype S and HVR-I haplotype XIV, both genotypes previously identified in all Asian *S. f. fuelleborni* isolates. The remaining positive sample was infected with three HVR-I haplotypes (GenBank accession numbers: PV489780, PV489781, PV489782): two with 275bp sequences identical to the only published sequence of *S. f. kellyi* (AJ417029) (13), and the third differing by a single SNP (T-C) at position 28. Based on this, we assigned this *Strongyloides* sp. to *S. f. kellyi*. HVR-I sequences of *S. f. kellyi* (432bp) differed from those of *S. ransomi* (LC324901, OP288111) by two SNPs and from *S. venezuelensis* (AB923887) by one SNP (Figure 4). At the HVR-IV locus, the 248bp sequence of *S. f. kellyi* was 100% identical to sequences of *S. ransomi* (OP288111, KU724127) and *S. venezuelensis* (AB923887).

**Figure 4.**
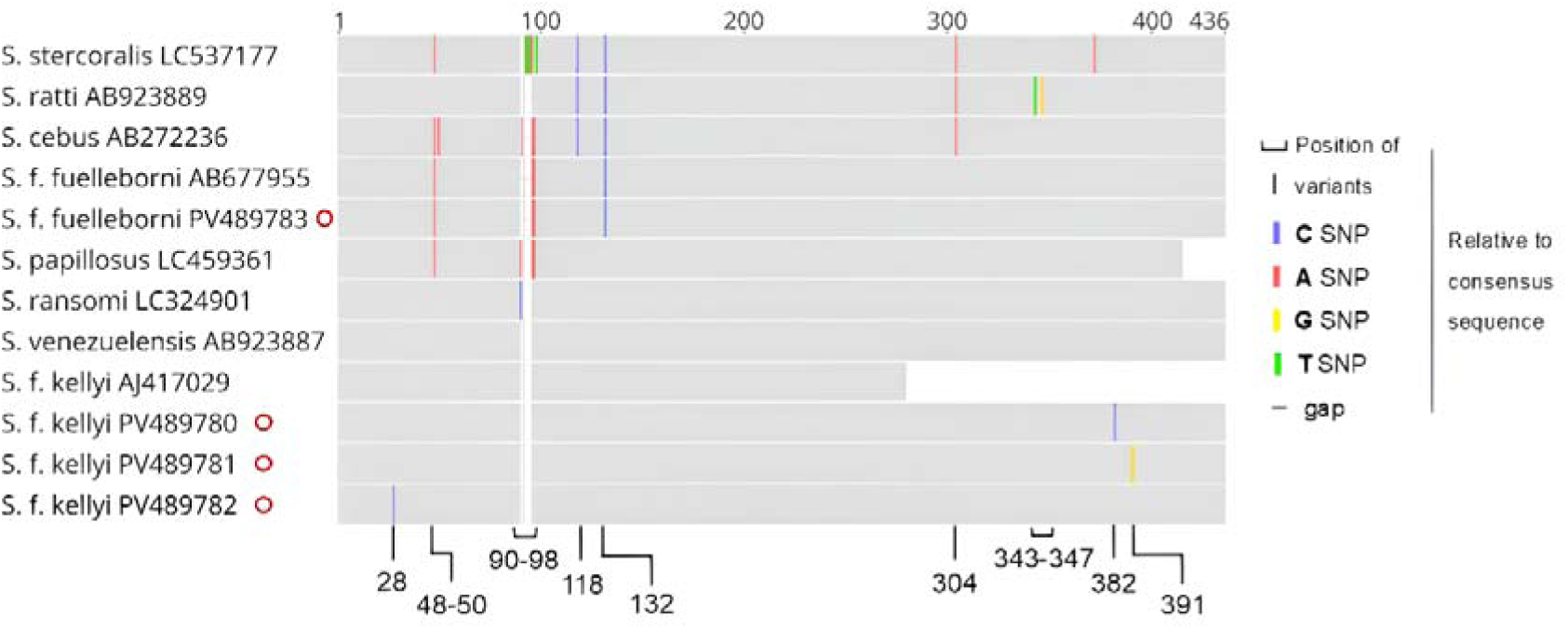
Schematic of MUSCLE-aligned *Strongyloides* spp. *18S rRNA* HVR-I sequences. Sequences obtained in this study are marked with red circles. Published sequences are annotated with their GenBank accession numbers.

*cox1* sequences were available for six samples, all assigned to *S. f. fuelleborni* (**Table 1**). The seventh sample, containing *S. f. kellyi*, did not amplify. Three separate *S. f. fuelleborni* haplotypes were identified (GenBank accession numbers: PQ774615, PQ774616, PQ774617). Maximum-likelihood and Bayesian inference phylogenetic analyses on the *cox1* locus placed PNG *S. f. fuelleborni* in a clade with *S. f. fuelleborni* from Myanmar *rhesus macaques* (OL672153). These sequences also clustered closely with *S. f. fuelleborni* from Bangladesh humans (OR805176 and OR805181) (**Figure 5**).

**Figure 5.**
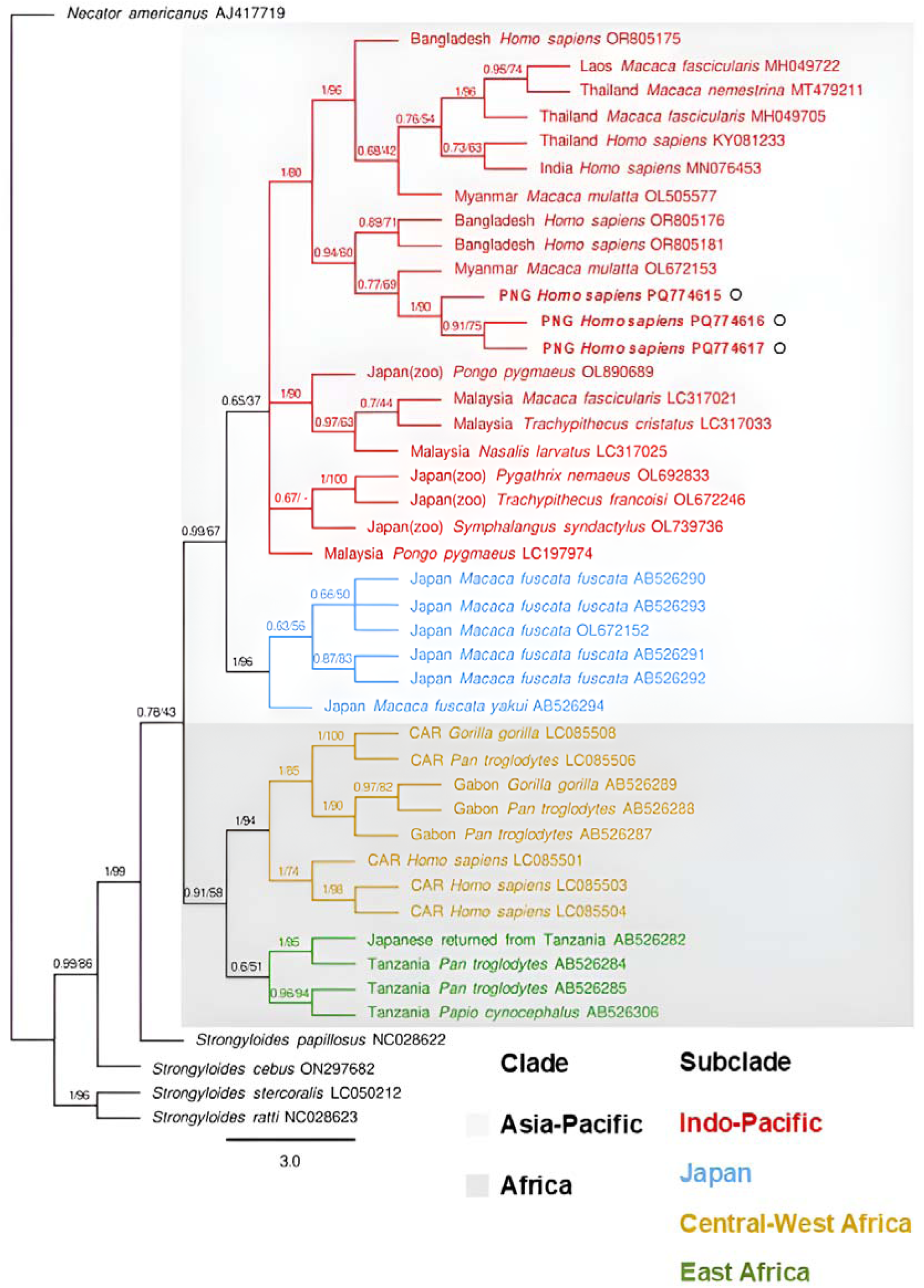
Phylogenetic tree of *Strongyloides fuelleborni fuelleborni* based on *cox*1 sequences. Bayesian posterior probability and maximum likelihood bootstrap support percentages (1000 replicates) are indicated at the nodes. Sequences obtained in this study are highlighted in bold and marked with black circles. Published sequences are annotated with their country of origin (CAR = Central African Republic), host species, and GenBank accession numbers. Clades and subclades of *S. f. fuelleborni* are colour-coded based on geographic regions.

## Discussion

Our study provides the first molecular evidence of the natural occurrence of *S. f. fuelleborni* in humans outside Asia and Africa. We also demonstrate that *S. f. kellyi* is taxonomically distinct from *S. f. fuelleborni*. For this reason, henceforth we consider the sub- species epithets *S. f. kellyi* and *S. f. fuelleborni* to be redundant and support elevation of *S. f. kellyi* to species status as “*Strongyloides kellyi*”, as previously suggested(2), once the requirements of the International Code of Zoological Nomenclature are fulfilled. Based on our findings, we hypothesise that at least three *Strongyloides* spp. infect humans in PNG: *S. stercoralis*, *S. f. fuelleborni*, and *S. kellyi*, and that high burden infections of the latter species being the aetiological agent of infantile SBS in New Guinea.

Viney et al. (1991) described very subtle morphological distinctions between *S.* cf *fuelleborni* from PNG humans and *S. f. fuelleborni* from African NHPs. Both nematodes differed markedly in morphology from *S. ransomi* found in PNG pigs. A separate isoenzyme electrophoretic analysis supported these findings. As these analyses did not include Asian isolates of *S. f. fuelleborni* and only a single egg-producing *Strongyloides* sp. was known to infect humans in PNG at that time, it remains possible that the New Guinea nematode examined by Viney et al. (1991) instead represented an Asian-Pacific strain of *S. f. fuelleborni*. If confirmed, this would support subspeciation for African and Asian-Pacific *S. f. fuelleborni*. Further genome-based phylogenetic analysis, combined with advanced morphological characterisation, of isolates from Africa, Asia, and the Pacific regions is needed to validate this hypothesis.

Our findings corroborate the work of Dorris et al. (2002), confirming that *S. kellyi* is likely a distinct species that is taxonomically more closely related to *S. ransomi* and *S. venezuelensis* than to *S. f. fuelleborni*. *S. f. kellyi* was indistinguishable from *S. ransomi* and *S. venezuelensis* at the *18S rRNA* HVR-IV locus and exhibited only 1-2 SNPs at the HVR-I locus, suggesting a recent common ancestry among these species. However, we cannot rule out the possibility that taxonomic positions inferred from such short sequences may be artifacts, and no mitochondrial sequences were available in this study to support these findings. Further phylogenetic analysis using whole genome or mitochondrial genome data is required for confirmation.

Our findings provide explanation for much of the unknown epidemiology of infant strongyloidiasis in PNG. Given that both *S. f. fuelleborni* and *S. f. kellyi* occur in PNG, previously reported epidemiologic data on “*S. f. kellyi*”, based on microscopic detection and identification of *Strongyloides* eggs, likely represent infections with either species. We observed a high incidence of *S. f. fuelleborni* infection (7/19) in PNG infants, consistent with historical reports of “*S. f. kellyi*” prevalence ranging from 20% to 93% in children, with rates reaching 60% within the first year of life (7–9, 14, 26). We found comparatively few *S. f. kellyi* infections (1/19), mirroring the similarly rare and sporadic occurrence of SBS previously reported in this population (7,9,10). Prior clinical, epidemiological, treatment, and biological data relating to *S. f. kellyi* and *S. f. kellyi* infections are likely confounded with *S. f. fuelleborni* infections, and therefore redundant. This work must now be revisited using species-specific molecular tools to differentiate these two agents.

While the pathogenesis of SBS remains enigmatic, the substantial genetic similarity between *S. f. kellyi* and *S. ransomi* observed in this study raises the possibility of a shared disease-causing mechanism between the two species. In newborn suckling piglets, *S. ransomi* infection is acquired from the mother via the transmammary route. It causes a protein losing enteropathy characterised by villus atrophy, malabsorption, diarrhoea, progressive dehydration, hypoproteinemia, anaemia, anorexia, emaciation, sudden death(34, 35) and reduced protein synthesis in the liver(36), a clinical picture strikingly similar to SBS in PNG infants (1, 7). Further research into the aetiology and epidemiology of SBS and its association with *S. f. kellyi* infection is important to understanding its public impact.

Equally important is understanding the transmission pattern of *S. f. fuelleborni*. In Africa and Asia, *S. f. fuelleborni* is a common infection of NHPs and is considered a zoonosis originating from these animals (24, 25). Both Pampiglione and Ricciardi (25) and Hira and Patel (26) suggested that human-to-human transmission of *S. f. fuelleborni* occurs in some regions of Africa. This was supported by genetic analysis by Barratt and Sapp (27), who identified a human-specific subpopulation among African *S. f. fuelleborni* isolates. Given the absence of an NHP reservoir on the island of New Guinea, it is likely that *S. f. fuelleborni* has adapted to exclusive human-to-human transmission after being introduced to PNG through human migration (28).

In this study, the youngest infant infected with *S. f. fuelleborni* was approximately three months old. Historical studies reported *Strongyloides* eggs in faeces of PNG infants as young as 18 days old (7). Sampling of breastmilk from mothers in PNG by Barnish and Ashford (28) did not identify any *Strongyloides* larvae; however, difficulties in obtaining faecal samples from these mothers left their infection status uncertain. In the same village, the prevalence of *Strongyloides* in adult faeces was only 14% (14). In a survey of 25 lactating mothers of infants with confirmed *S. f. fuelleborni* infection in the Democratic Republic of Congo, *Strongyloides* autoinfective L3 larvae were identified in the breastmilk of one mother(29). This suggested that transmammary transmission could be responsible for the high infection rates in infants as young as 50-74 days in that region(29). Given that our genetic analysis indicates that these worms belong to clades of the same species, it is plausible that transmammary transmission to PNG infants may occur. This speculation warrants further investigation, and the use of molecular genotyping tools may be necessary to track transmission pattern.

We note that not all diagnostic *Strongyloides* qPCR assays detect *S. f. fuelleborni* infection. The Llewelyn modification(20) of Verweij’s(30) qPCR appears generic and detected all except one *S. f. fuelleborni* infection. Conversely, the Pilotte *S. stercoralis* qPCR(21) only amplified *S. kellyi* DNA, while not detecting *S. f. fuelleborni* infections. This demonstrates that in future surveillance of *Strongyloides* spp. infections in humans and animals, the specificity of PCR diagnostics employed must be considered, as the choice of qPCR may markedly impact the findings of any survey.

## Conclusions

We present the first molecular evidence of *S. f. fuelleborni* infection in humans in the Pacific. Our findings demonstrate that at least two egg-producing *Strongyloides* spp. infect humans in PNG (and by extension potentially the entire island of New Guinea). The occurrence of *S. f. fuelleborni* and incapacity to morphologically differentiate these eggs from those of *S. f. kellyi* explains previous confusion regarding the aetiology of SBS. We also demonstrated that *S. f. kellyi* is taxonomically distinct from *S. f. fuelleborni.* It is hypothesised that high burden infection with *S. f. kellyi*, a species closely related to *S. ransomi* in pigs, is responsible for infantile SBS in New Guinea. Further studies on these two helminth infections in New Guinea are warranted and should employ molecular tools to capable of differentiating the two species.

## Data Availability

All data produced in the present work are contained in the manuscript

## Acknowledgements

We thank the participants and their families for providing stool samples. We thank the leadership and staff at Papua New Guinea Institute of Medical Research, particularly members of the Infection and Immunity Unit, for their collaboration on this project. This work was partially funded by the National Health and Medical Research Council (NHMRC) Investigator Grant APP1194462. HZ and JH receive an Australian Government Research Training Program Scholarship from James Cook University, Australia.

## Biographical Sketch

Ms Zhao is a PhD candidate at James Cook University. Her research focuses on the taxonomy and molecular epidemiology of *Strongyloides* species in humans and companion animals.

